# *De novo* variants in the non-coding spliceosomal snRNA gene *RNU4-2* are a frequent cause of syndromic neurodevelopmental disorders

**DOI:** 10.1101/2024.04.07.24305438

**Authors:** Yuyang Chen, Ruebena Dawes, Hyung Chul Kim, Sarah L Stenton, Susan Walker, Alicia Ljungdahl, Jenny Lord, Vijay S Ganesh, Jialan Ma, Alexandra C Martin-Geary, Gabrielle Lemire, Elston N D’Souza, Shan Dong, Jamie M Ellingford, David R Adams, Kirsten Allan, Madhura Bakshi, Erin E Baldwin, Seth I Berger, Jonathan A Bernstein, Natasha J Brown, Lindsay C Burrage, Kimberly Chapman, Alison G Compton, Chloe A Cunningham, Precilla D’Souza, Emmanuèle C Délot, Kerith-Rae Dias, Ellen R Elias, Carey-Anne Evans, Lisa Ewans, Kimberly Ezell, Jamie L Fraser, Lyndon Gallacher, Casie A Genetti, Christina L Grant, Tobias Haack, Alma Kuechler, Seema R Lalani, Elsa Leitão, Anna Le Fevre, Richard J Leventer, Jan E Liebelt, Paul J Lockhart, Alan S Ma, Ellen F Macnamara, Taylor M Maurer, Hector R Mendez, Stephen B Montgomery, Marie-Cécile Nassogne, Serena Neumann, Melanie O’Leary, Elizabeth E Palmer, John Phillips, Georgia Pitsava, Ryan Pysar, Heidi L Rehm, Chloe M Reuter, Nicole Revencu, Angelika Riess, Rocio Rius, Lance Rodan, Tony Roscioli, Jill A Rosenfeld, Rani Sachdev, Cas Simons, Sanjay M Sisodiya, Penny Snell, Laura St Clair, Zornitza Stark, Tiong Yang Tan, Natalie B Tan, Suzanna EL Temple, David R Thorburn, Cynthia J Tifft, Eloise Uebergang, Grace E VanNoy, Eric Vilain, David H Viskochil, Laura Wedd, Matthew T Wheeler, Susan M White, Monica Wojcik, Lynne A Wolfe, Zoe Wolfenson, Changrui Xiao, David Zocche, John L Rubenstein, Eirene Markenscoff-Papadimitriou, Sebastian M Fica, Diana Baralle, Christel Depienne, Daniel G MacArthur, Joanna MM Howson, Stephan J Sanders, Anne O’Donnell-Luria, Nicola Whiffin

## Abstract

Around 60% of individuals with neurodevelopmental disorders (NDD) remain undiagnosed after comprehensive genetic testing, primarily of protein-coding genes^1^. Increasingly, large genome-sequenced cohorts are improving our ability to discover new diagnoses in the non-coding genome. Here, we identify the non-coding RNA *RNU4-2* as a novel syndromic NDD gene. *RNU4-2* encodes the U4 small nuclear RNA (snRNA), which is a critical component of the U4/U6.U5 tri-snRNP complex of the major spliceosome^2^. We identify an 18 bp region of *RNU4-2* mapping to two structural elements in the U4/U6 snRNA duplex (the T-loop and Stem III) that is severely depleted of variation in the general population, but in which we identify heterozygous variants in 119 individuals with NDD. The vast majority of individuals (77.3%) have the same highly recurrent single base-pair insertion (n.64_65insT). We estimate that variants in this region explain 0.41% of individuals with NDD. We demonstrate that *RNU4-2* is highly expressed in the developing human brain, in contrast to its contiguous counterpart *RNU4-1* and other U4 homologs, supporting *RNU4-2*’s role as the primary U4 transcript in the brain. Overall, this work underscores the importance of non-coding genes in rare disorders. It will provide a diagnosis to thousands of individuals with NDD worldwide and pave the way for the development of effective treatments for these individuals.

## Main

Despite increasingly powerful genomic and analytic approaches for the diagnosis of rare developmental disorders, currently ∼60% of individuals remain without an identified genetic diagnosis after genomic testing with current methods^1^. To date, the overwhelming majority of known disease-causing variants are in the ∼1.5% of the genome that directly encodes proteins^3^. In contrast, the non-coding genome (that makes up the remaining 98.5%) has been relatively unexplored, especially regions far from protein-coding genes. Large-scale, systematic application of genome sequencing to clinical populations has increasingly enabled investigation of the contribution of variants in non-coding regions to genetic disorders^4^.

Non-coding RNAs, which comprise 37.4% of processed exonic RNA sequence in humans^5^, include important regulators of biological processes with diverse roles across cells and tissues^6^. Small nuclear RNAs (snRNAs) are a subcategory of non-coding RNAs that are key components of the spliceosome^7^. snRNAs complex with a multitude of proteins and other snRNA species in small nuclear ribonucleoprotein (snRNP) complexes to mediate the removal of introns from pre-mRNA transcripts^8^. Many spliceosome components have a demonstrated role in human disorders, including two snRNA components of the minor spliceosome: *RNU12* variants cause autosomal recessive early-onset cerebellar ataxia^9^, while *RNU4ATAC* variants cause an autosomal recessive multisystem congenital disorder including microcephaly, growth retardation, and developmental delay (eponyms include Taybi Linder^10^, Lowry-Wood^11^ and Roifman syndromes^12^).

Here, we identify variants in *RNU4-2*, which encodes the U4 snRNA component of the major spliceosome, as a newly recognised autosomal dominant disorder. Using a cohort of 8,841 probands with genetically undiagnosed NDD in Genomics England (GEL)^4^, we identify variants in a critical 18 base-pair (bp) region in the centre of *RNU4-2* associated with a severe neurodevelopmental phenotype and estimate that variants in this region explain ∼0.41% of individuals with neurodevelopmental disorders (NDD). We demonstrate that variants in this region are severely depleted from large population datasets. We show that NDD variants map to critical structural elements in the U4/U6 complex that are important to correctly position U6 ACAGAGA to receive the 5’ splice-site during initial spliceosome activation, and detail the expression of *RNU4-2* through brain development.

### A highly recurrent insertion explains 0.52% of undiagnosed NDD in Genomics England

We identified a highly recurrent single base insertion (GRCh38:chr12:120,291,839:T:TA; n.64_65insT) in *RNU4-2* in GEL^1^. This variant was initially identified as arising *de novo* in 38 probands recruited for genome sequencing with their unaffected parents^13^. Extending the search to include probands without data for both parents in the full GEL cohort, we identified an additional eight individuals with the n.64_65insT variant; in all eight, the detectable inheritance is consistent with the variant having arisen *de novo* (i.e. where a single parent sample was available the variant was not detected in it). All of the 46 individuals with the variant have undiagnosed NDD (categorised as global developmental delay, intellectual disability, and/or autism spectrum disorder), corresponding to 0.52% of 8,841 probands with currently undiagnosed NDD in GEL. The n.64_65insT variant is not found in any of 3,408 NDD probands with an existing genetic diagnosis, 21,817 probands with non-NDD phenotypes, or in 33,122 unaffected individuals. Individuals with the variant are significantly enriched for global developmental delay (n=37; OR=3.56; Fisher’s *P*=2.75×10^-4^), delayed gross motor development (n=26; OR=2.55; *P*=1.64×10^-3^), microcephaly (n=26; OR=6.62; *P*=7.87×10^-10^), delayed fine motor development (n=24; OR=2.61; *P*=1.69×10^-3^), hypotonia (n=18; OR=3.60; *P*=7.09×10^-5^), short stature (n=15; OR=3.54; *P*=2.17×10^-4^), drooling (n=7; OR=19.2; *P*=2.83×10^-7^), and absent speech (n=6; OR=6.23; *P*=7.45×10^-4^) compared to all other probands with NDD in GEL (n=12,203; diagnosed and undiagnosed) (**Figure 1A; Supplementary Table 1**).

**Figure 1:**
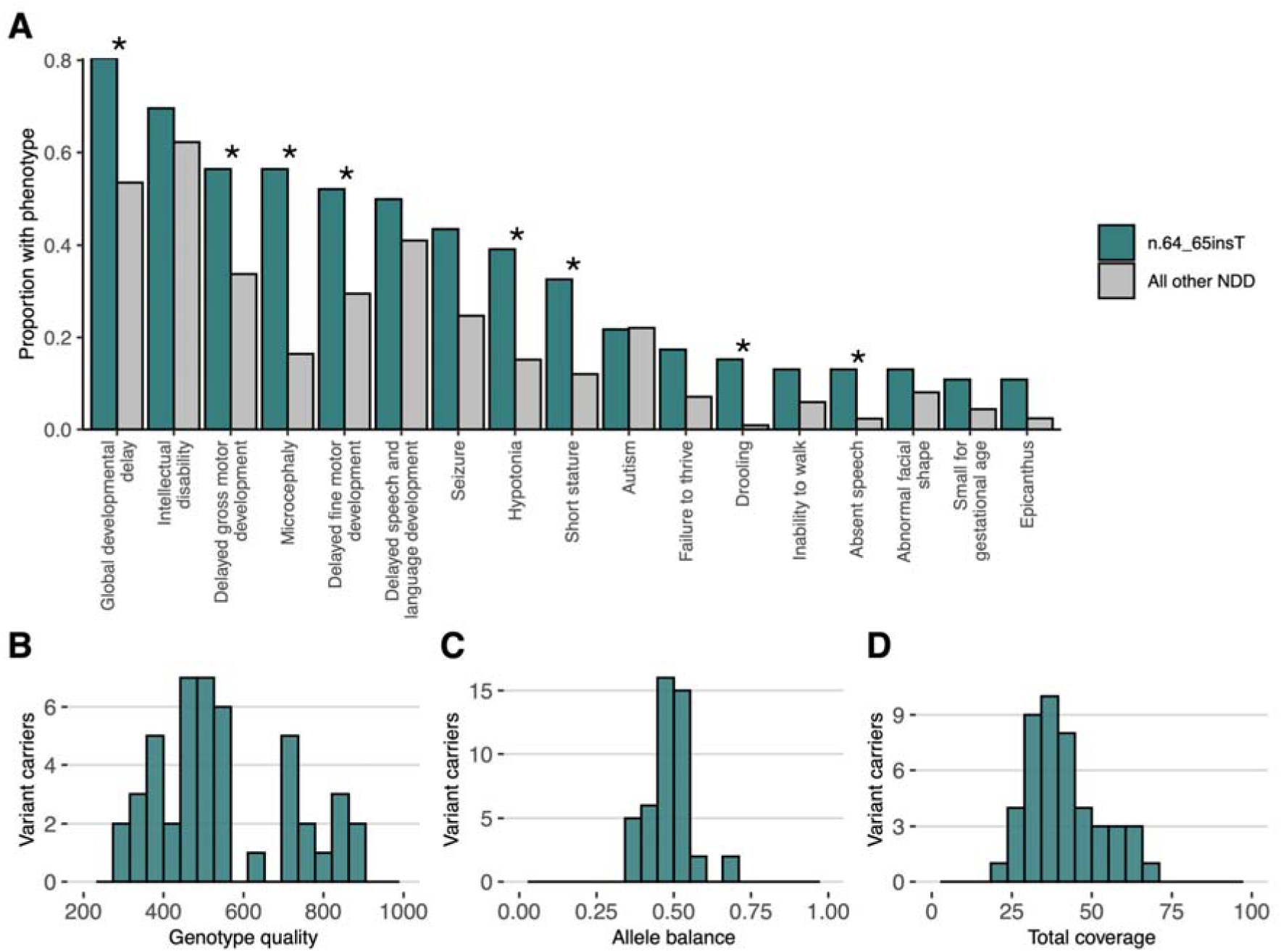
Characterisation of individuals with the n.64_65insT variant in GEL. (A) The proportion of individuals with human phenotype ontology (HPO) terms corresponding to phenotypes observed in ≥ 5 individuals with the n.64_65insT variant compared to all other individuals with NDD. Terms that are significantly enriched in individuals with the n.64_65insT variant are marked with a *. Multiple terms relating to global developmental delay, intellectual disability, hypotonia, seizure, microcephaly, autism, and short stature have been collapsed into single phenotypes. Of note, this figure relates only to HPO terms entered for each individual into GEL, which may be incomplete. A more detailed phenotypic characterisation of individuals with variants in *RNU4-2* is provided below. (B-D) Quality control metrics for the variant calls in all 46 individuals with the variant: (B) genotype quality scores, (C) allele balance, and (D) coverage.

The n.64_65insT variant is not found in 76,215 genome-sequenced individuals in gnomADv4.0^14^, or in 245,400 individuals in the All of Us dataset^15^. It is seen in a single individual in the UK Biobank^16^ (allele frequency=1.02×10^-6^) with a variant allele balance consistent with a true variant (23 reference and 18 [44%] alternate reads). This individual has an ICD-10 code for ‘personal history of disease of the nervous system and sense organs’ but no further phenotype data to assess a potential NDD diagnosis (**Supplementary Table 2)**.

Given the high recurrence rate of this insertion, we wanted to rule out that it is a sequencing or mapping error, despite the overwhelming evidence of phenotype enrichment. Notably, the variant is a single A insertion after a run of four Ts, ruling out the most common cause of sequencing error for indels, polymerase slippage in homopolymer repeats. The variant calls were all high quality based on both analysis of quality metrics (**Figure 1B-D**) and manual inspection on IGV (**Supplementary Figure 1**). Finally, the genomic region surrounding the insertion and *RNU4-2* maps uniquely to a single region of the genome with short-read sequencing in GRCh38 and T2T CHM13v2.0/hs1 (**Supplementary Figure 2**).

### The n.64_65insT variant is within a highly constrained region with multiple NDD-causing variants

The recurrent n.64_65insT variant resides within the central region of *RNU4-2*, towards the 5’ end of an 18 bp region which is depleted of variants in population datasets compared with the rest of the gene (26% of all possible SNVs observed in UK Biobank compared to a median of 78% across the rest of the gene; **Figure 2A; Supplementary Figure 3**). Based on the population variant data, we defined a critical, highly constrained region as chr12:120,291,825-120,291,842.

**Figure 2:**
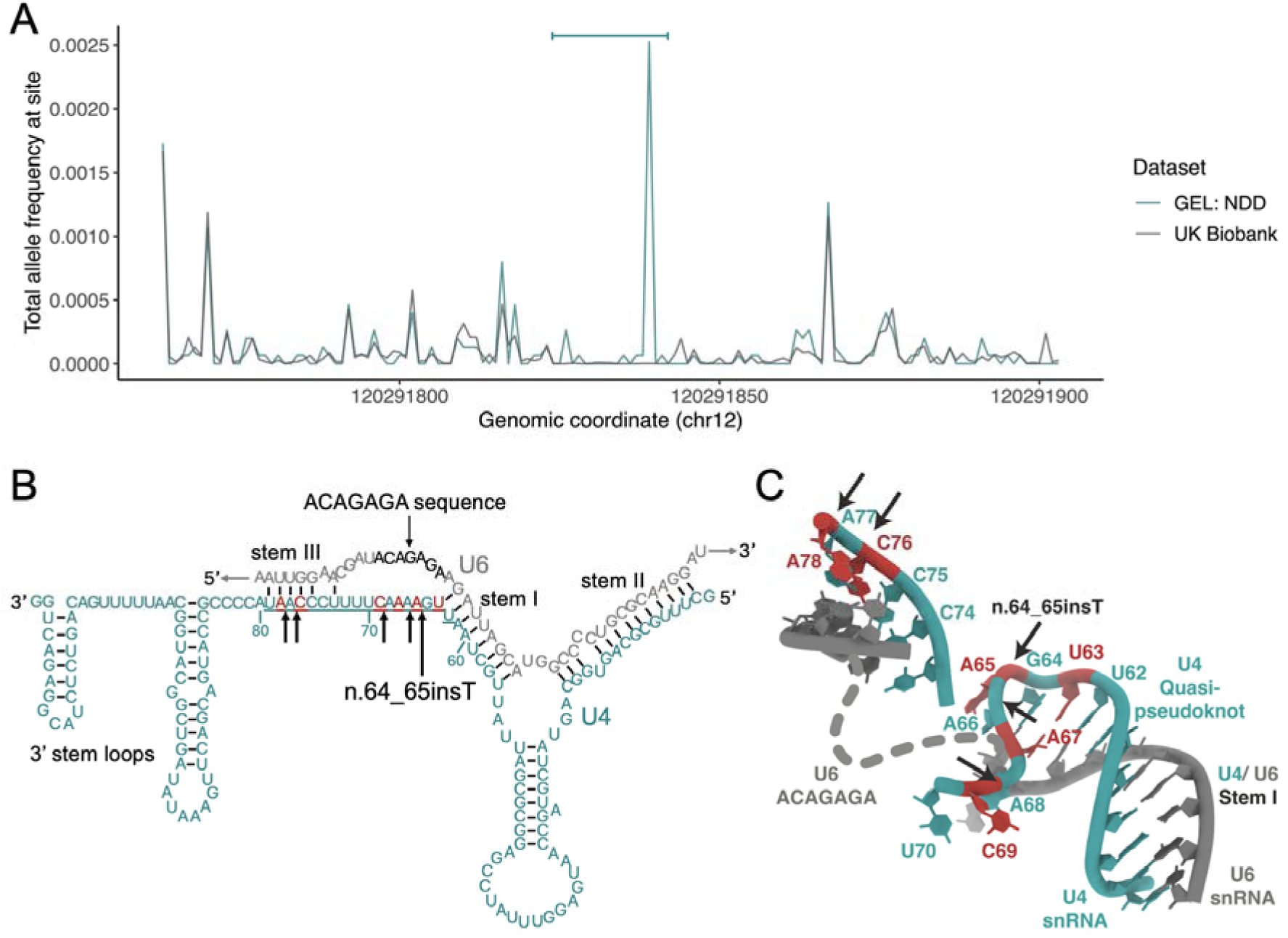
A highly structured 18 bp region of RNU4-2 that is critical for BRR2 helicase activity is enriched for variants in NDD and depleted in population cohorts. (A) Allele frequency of variants in 7,519 undiagnosed NDD probands GEL (teal) and the UK Biobank cohort (grey) across *RNU4-2*. The 18 bp critical region is marked by a horizontal bar at the top of the plot. (B) Schematic of U4 (teal) binding to U6 snRNA (grey). The 18 bp critical region is underlined. (C) The structure of U4 and U6 snRNAs resolved by cryoEM^21^. Created using RCSB Protein Data Bank^22^ (structure 6QW6). In both (B) and (C) single base insertions identified in individuals with NDD are shown by black arrows and positions of SNVs by red nucleotides.

We searched for variants across this region in GEL, and also in additional cohorts containing undiagnosed individuals with NDD (see **methods**). In total, we identified 119 individuals with variants across this region (**Table 1**), the vast majority of which have the initial n.64_65insT variant (n=92; 77.3%). For 92 of the 119 individuals, sequencing data for both parents was available to confirm the variants had arisen *de novo*. Five of the 11 additional variants are also single base insertions, including n.77_78insT (GRCh38:chr12:120,291,826:T:TA), which is seen in six individuals, two of whom are affected siblings. The enrichment of single base insertion variants in this region in individuals with NDD is striking: 54/8,841 (0.61%) GEL undiagnosed NDD probands (55/10,388 individuals) have single base insertions compared to 2/490,132 individuals in the UK Biobank (OR=1,531; 95%CI:404,>16,384; Fisher’s *P*=3.3×10^-92^).

**Table 1:** Variants identified in individuals with NDD in the 18 bp critical region of *RNU4-2*. (chr12:120,291,825-120,291,842). Numbers in brackets in NDD count columns correspond to individuals with detailed clinical information in **Table 2**. The count in population cohorts is shown only for variants observed in individuals with NDD. A full list of variants found across the region in population cohorts is in **Supplementary Table 3**. *count includes two siblings. **NHS GMS (n=21); MSSNG^17^ (n=2); SSC^18^ (n=1); GREGoR (n=10); Undiagnosed Diseases Network^19^ (UDN; n=8); from personal communication/Matchmaker Exchange (n=17).

| variant | nucleotide description | GEL NDD count (in Table 2) | Non-GEL NDD count** (in Table 2) | population cohort count |
| --- | --- | --- | --- | --- |
| **Single base insertions** |  |  |  |  |
| 12:120291839:T:TA | n.64_65insT | 46 (2) | 46 (31) | 1 (UK Biobank) |
| 12:120291839:T:TC | n.64_65insG | 0 | 2 (1) | 0 |
| 12:120291826:T:TA | n.77_78insT | 6* | 0 | 0 |
| 12:120291827:T:TA | n.76_77insT | 1 | 0 | 0 |
| 12:120291835:G:GT | n.68_69insA | 1 | 0 | 0 |
| 12:120291838:T:TA | n.65_66insT | 1 | 0 | 0 |
|  | **Total** | **55\*** | **48** | **1** |
| **SNVs** |  |  |  |  |
| 12:120291839:T:C | n.65A>G | 2 | 0 | 0 |
| 12:120291826:T:G | n.78A>C | 1 | 0 | 0 |
| 12:120291828:G:A | n.76C>T | 1 | 6 (1) | 1 (gnomAD v4) |
| 12:120291835:G:A | n.69C>T | 0 | 1 (1) | 0 |
| 12:120291837:T:C | n.67A>G | 1 | 3 | 0 |
| 12:120291841:A:C | n.63T>G | 1 | 0 | 0 |
|  | **Total** | **6** | **10** | **1** |

Aside from insertions, there is also a modest enrichment of SNVs in GEL NDD probands across the critical region (undiagnosed NDD: 6/8,841; UK Biobank 35/490,132; OR=9.51; 95%CI:3.27-22.8; Fisher’s *P*=8.16×10^-5^). We identified 16 individuals across cohorts with SNVs in this region (**Table 1**; 11 confirmed *de novo*), all with phenotypes consistent with individuals with insertion variants. The identified SNVs cluster with the two regions harbouring insertion variants at the extreme ends of the 18 bp critical region (**Figure 2B**). Conversely, SNVs in the central portion (particularly at nucleotides 71-74) are observed in both non-NDD individuals in GEL (n=2) and population controls, although all at low frequencies (**Supplementary Table 3**). Across the remainder of *RNU4-2* there is no significant enrichment of variants in undiagnosed NDD probands when compared to non-NDD probands (194/7,519 undiagnosed NDD; 521/19,428 non-NDD in GEL aggregated variant dataset^20^; OR=0.96; 95%CI:0.81-1.14; Fisher’s P=0.67).

In total, we identify variants in this 18 bp region in 119 individuals with NDD. This includes 60/8,841, or 0.68%, of all genetically undiagnosed NDD probands in GEL (0.49% of all NDD probands). In contrast, variants in this region are observed in 39/490,132 (0.008%) individuals in the UK Biobank (OR=85.8; 95%CI:56.4-131.6; Fisher’s P=1.84×10^-78^).

U4 snRNA binds to U6 snRNA through extensive complementary base-pairing in the U4/U6.U5 tri-snRNP complex of the major spliceosome. Unwinding of U4 and U6 is essential to generate the catalytically active spliceosome^2^. The 18 bp critical region in *RNU4-2* maps to a single-stranded region of U4 between the stem I region of complementary base-pairing to U6 and the 3’ stem-loop structures (nucleotides 62 to 79; **Figure 2B**). This region is known to be loaded into the active site of the *SNRNP200*-encoded BRR2 helicase, which mediates unwinding of the U4/U6 duplex^2^. The highly recurrent n.64_65insT variant is within a previously described ‘quasi pseudoknot’, or T-loop, structure^21^ (**Figure 2C**). The region spanning nucleotides 76 to 78, where the recurrent n.77_78insT variant resides, is involved in base-pairing with U6 in stem III^23^ (**Figure 2C**). Both of these regions are thought to stabilise the U4/U6 interaction and accurately position the U6 ACAGAGA sequence to receive the 5’splice site during spliceosome activation. Insertion of a single base into either of these structures may destabilise the U4/U6 interaction and/or alter the positioning of the U6 ACAGAGA sequence and potentially disrupt the correct loading of the 5’ splice site into the fully assembled spliceosome. Nearby regions that are predicted to have important roles, such as the U4/U6 stem I binding region, are not enriched for variants in NDD probands.

### Variants in this crucial region cause a severe syndromic NDD phenotype

To characterise the phenotypic spectrum associated with variants in *RNU4-2*, we collected detailed phenotypic information for a subset of 36 individuals (33 with n.64_65insT, one with n.64_65insG, and two with SNVs; **Table 2**; **Supplementary Table 4**). Using these data, we find the *RNU4-2* syndromic NDD to be characterised by moderate to severe global developmental delay (two children with SNVs with moderate delay) and intellectual disability in all individuals. The majority (82%) achieved ambulation but at a delayed age (average 3.6 years, range 18 months to 7.5 years) with many noted to have a wide-based or ataxic gait. Only one individual (with an SNV) had fluent speech, some had a few words, and most were non-verbal. All but one were reported to have dysmorphic facial features. These facial features varied but consisted of a myopathic face with deep set eyes (some widely spaced and some narrowly spaced), epicanthus, wide nasal bridge, anteverted nares or underdeveloped ala nasi coll, large cupped ears (some posteriorly rotated), full cheeks, a distinctive mouth with full lips with downturned corners, high arched palate, and a large or protruding tongue.

**Table 2:**
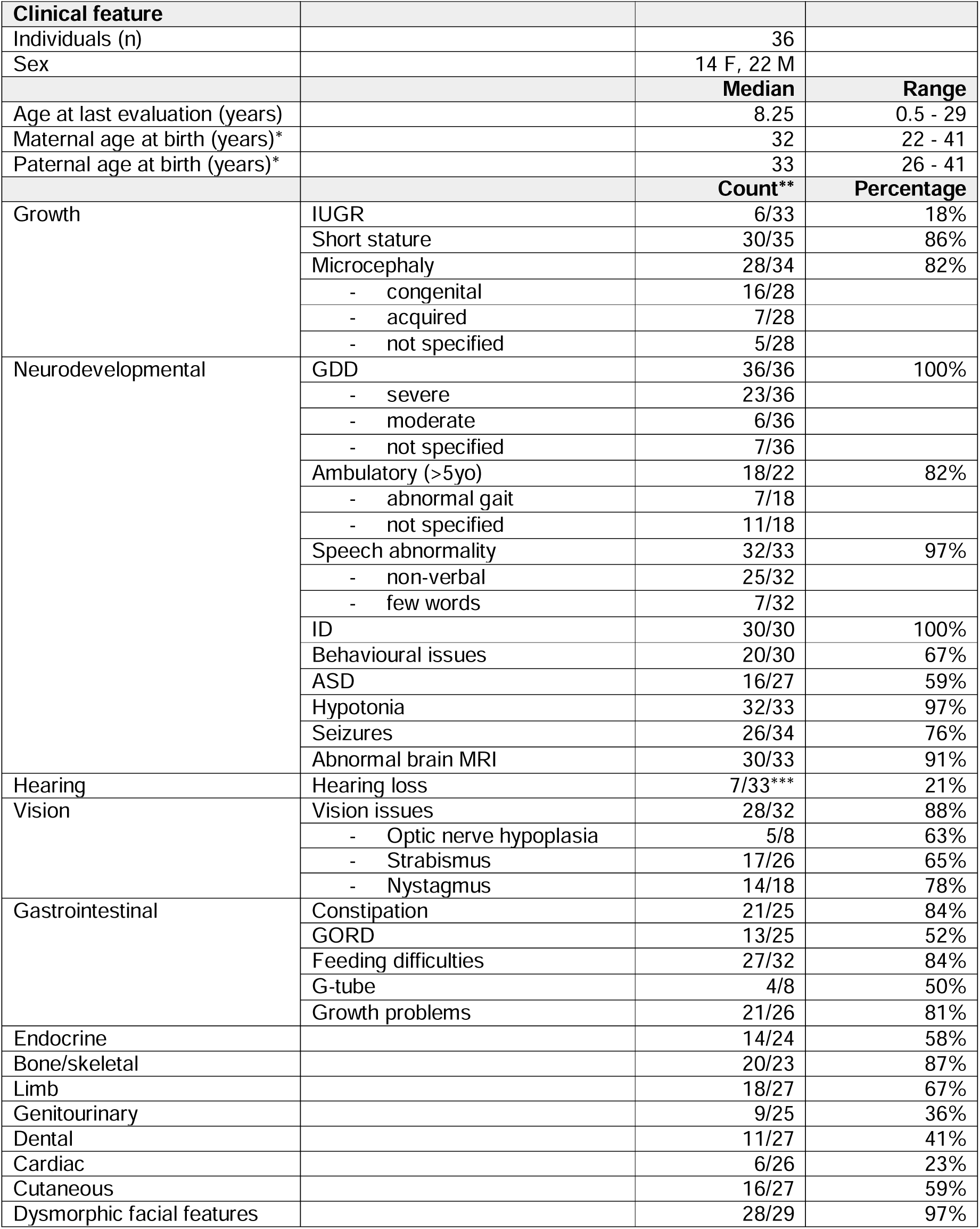
Clinical features of 36 individuals with *RNU4-2* variants. F, female; M, male; IUGR, intrauterine growth restriction; GDD, global developmental delay; ID, intellectual disability; ASD, autism spectrum disorder; MRI, magnetic resonance imaging; GORD, gastro-oesophageal reflux disease; GH, growth hormone; G-tube, gastrostomy tube *parental age only available for 27/36 individuals **denominator indicates the number of individuals for whom data were available ***one individual has a dual diagnosis in *GJB2* which would account for the hearing loss

Associated growth and neurodevelopmental phenotypes present in ≥75% of individuals include short stature, microcephaly (mostly congenital), seizures (spanning infantile spasms, focal seizures and generalised tonic-clonic seizures, febrile seizures, and status epilepticus with variable onset from the first year of life, but most between 3-10 years of age), and hypotonia. Brain MRI showed a spectrum of abnormalities in the majority of individuals, most frequently reduced white matter volume, non-specific abnormalities of the white matter, hypoplasia of the corpus callosum, ventriculomegaly, and delayed myelination. Involvement of multiple organ systems was reported for all individuals, often including visual (optic nerve hypoplasia, cortical blindness, strabismus, nystagmus), gastrointestinal (constipation, reflux, feeding issues with need for a gastrostomy tube), and bone/skeletal abnormalities (osteopenia, recurrent fractures, scoliosis, kyphosis, hip dysplasia), and in a lesser number of individuals, hearing, endocrine (hypothyroidism, growth hormone deficiency), limb, sleep, genitourinary, dental, cardiac, and cutaneous concerns (**Table 2; Supplementary Table 4**).

### RNA-sequencing in blood does not show a global disruption to splicing

Given the importance of U4 snRNA in the spliceosome and previous observations of global disruption to splicing observed in other spliceosomopathies^24^, we analysed RNA sequencing data from blood samples for five individuals from GEL. Three of these individuals have the highly recurrent n.64_65insT variant, another has the other recurrent insertion, n.77_78insT, and the final patient has an SNV (n.78A>C). We did not see any significant difference in the number of gene expression outliers using OUTRIDER^25^, or in the number of retained introns, or all outlier events using FRASER2^26^ in the five individuals with *RNU4-2* variants compared with 5,409 controls (**Supplementary Table 5**). At present, RNA from additional tissues (e.g. brain samples) of affected individuals is not available. It is possible that the observed *RNU4-2* variants disrupt more subtle aspects of alternative splicing in a tissue-specific manner, as has been observed for other snRNA variants^27^.

### RNU4-2 is highly expressed across tissues and in the brain across development

Humans have multiple genes that encode the U4 snRNA, although only two of these, *RNU4-2* and *RNU4-1,* are highly expressed in the human brain (**Supplementary Table 6**). *RNU4-2* and *RNU4-1* are contiguous on chr12, both 141 bp long, and highly homologous, differing by four nucleotides (97.2% homology). *RNU4-1* has a similar depletion of variants in population cohorts in the centre of the RNA, however, we do not observe an enrichment of variants in GEL in this central region (**Supplementary Figure 4**). There is a variant equivalent to our highly recurrent variant in *RNU4-1* that is observed in six individuals in the UK Biobank dataset. There are no consistent phenotypes recorded in these six individuals (**Supplementary Table 2**).

To investigate the reason for variants in *RNU4-2*, but not *RNU4-1*, causing NDD, we analysed the expression of both *RNU4-1* and *RNU4-2* in the brain. First, we analysed the expression patterns of both genes across multiple developmental stages using bulk RNA-seq data from 176 human prefrontal cortex samples in BrainVar^28^. The expression of *RNU4-1* and *RNU4-2* is tightly correlated (**Supplementary Figure 5**), however, *RNU4-2* is consistently expressed at a significantly higher level than *RNU4-1* (**Figure 3A**). Secondly, we assessed chromatin accessibility in the chromosome 12 locus containing both *RNU4-1* and *RNU4-2* using ATAC-seq data from two human prenatal prefrontal cortex samples. These data show a dramatic chromatin accessibility signal around *RNU4-2* and a much lower signal surrounding *RNU4-1*, again consistent with much higher expression of *RNU4-2* in the brain (**Figure 3B**). Overall, these data support the role of *RNU4-2* as the major U4 transcript in the brain.

**Figure 3:**
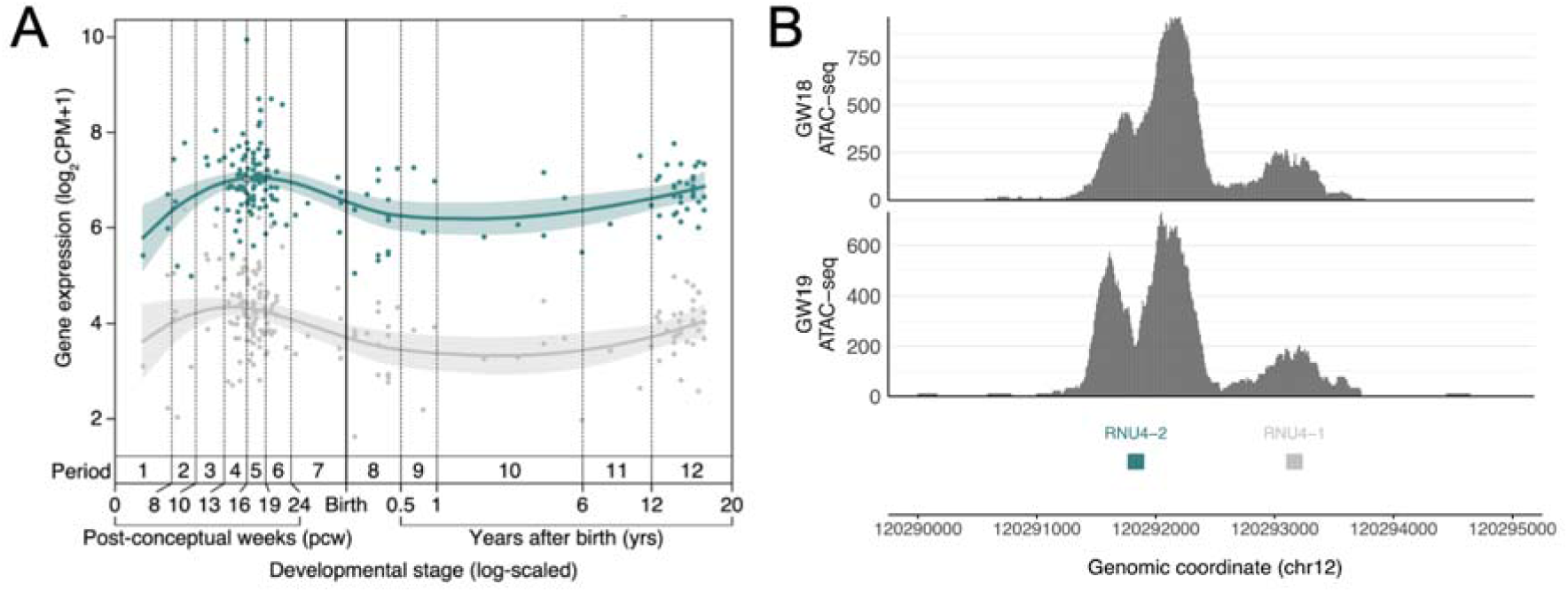
*RNU4-2* is more highly expressed than *RNU4-1* in the prefrontal cortex. (A) Levels of *RNU4-1* (grey) and *RNU4-2* (teal) expression at different developmental stages from BrainVar^28^. (B) ATAC-seq data from human prenatal prefrontal cortex (18 and 19 gestational weeks (GW)) with substantially higher peaks of chromatin accessibility around *RNU4-2* (teal) than *RNU4-1* (grey).

### Multiple factors likely explain the high recurrence of the n.64_65insT variant

The n.64_65insT variant is highly recurrent. It is observed in 46/12,249 NDD probands in GEL (0.38%; or 0.52% of undiagnosed NDD probands). In contrast, the most recurrent protein-coding variant in a dataset of 31,058 individuals with developmental disorders^29^ is observed in 36 individuals (0.12%; GRCh38:chr11:66211206:C:T; PACS1:p.Arg203Trp). The exact reasons for this high recurrence are unclear, however, we hypothesise three contributing factors. First, a high local mutation rate, which may be driven by the open chromatin state and very high levels of transcription (**Figure 3**). In UK Biobank, a median of 76% of all possible SNVs in *RNU4-2* are observed (calculated across 18 bp sliding windows). This is compared with 13% on average in 1,000 random intergenic sequences of the same length (141 bp; *P*<0.001, Monte-Carlo Fisher-Pitman test; **Supplementary Figure 6**). Despite the high number of variants in *RNU4-2* in UK Biobank, there are no individuals with homozygous variants and all observed variants are very rare (maximum allele frequency = 0.025%), consistent with high levels of selection acting on variants across *RNU4-2*.

Secondly, a high overall mutational burden does not explain the high recurrence of this specific single base insertion. Local formation of secondary structure and base stacking is a known driver of biased small insertion mutations^30^. The high propensity of this region to form secondary structure when single-stranded may drive creation of this specific insertion. Finally, it may be that germline selection is acting to increase the frequency of this specific variant, as has been shown for other highly recurrent sites^31^. While we see no association with paternal age (mean 33.1 in probands with *RNU4-2* variants and 33.4 across other NDD probands; **Supplementary Figure 7**), fully testing this hypothesis will require deep sequencing of testes or sperm samples.

### No other spliceosomal snRNA genes are enriched for de novo variants in NDD

Given the newly identified importance of *RNU4-2* in NDD, we sought to determine whether other snRNA genes with no known association to NDD could also harbour novel diagnoses. We investigated 28 snRNA genes that are expressed in the brain, using multiple approaches (**Supplementary Table 7**). First, we tested for an overall enrichment of *de novo* variants in undiagnosed NDD probands compared to non-NDD probands across each snRNA with at least two identified *de novo* variants in probands with undiagnosed NDD (n=13) using the high-confidence *de novo* callset in GEL. Of the 12 genes other than *RNU4-2*, none showed a significant enrichment of *de novo* variants in undiagnosed NDD probands (all Fisher’s *P*>0.15).

Secondly, hypothesising that the burden of pathogenic variants in other snRNAs may be restricted to specific critical regions, as we see for *RNU4-2*, we used an 18 bp sliding window to identify snRNA regions that are depleted of variation in the UK Biobank compared to the overall variant burden across each gene. Notably, the regions with the highest depletion in *RNU4ATAC* correspond to two hotspots of pathogenic variants in ClinVar (chr2:121530923-121530946, chr2:121530984-121531007), however, the strength of the depletion in these regions is lower than in *RNU4-2 (minimum normalised proportion of observed −0.11 and −0.2 versus −0.5 for the depleted region in RNU4-2)*, consistent with lower selection acting on variants in *RNU4ATAC* that cause recessive disorders. We identified 14 regions in 13 unique snRNAs with a deviation from the median number of SNVs across the full gene of at least 20% (**Figure 4**; **Supplementary Table 8**). We repeated our *de novo* variant enrichment test in regions with at least two *de novo* variants in undiagnosed NDD probands (n=3). Only the conserved region in *RNU4-2* was significant (Fisher’s P=1.34×10^-9^; undiagnosed NDD probands n=33, non-NDD probands n=0; all other tests Fisher’s *P*>0.25).

**Figure 4:**
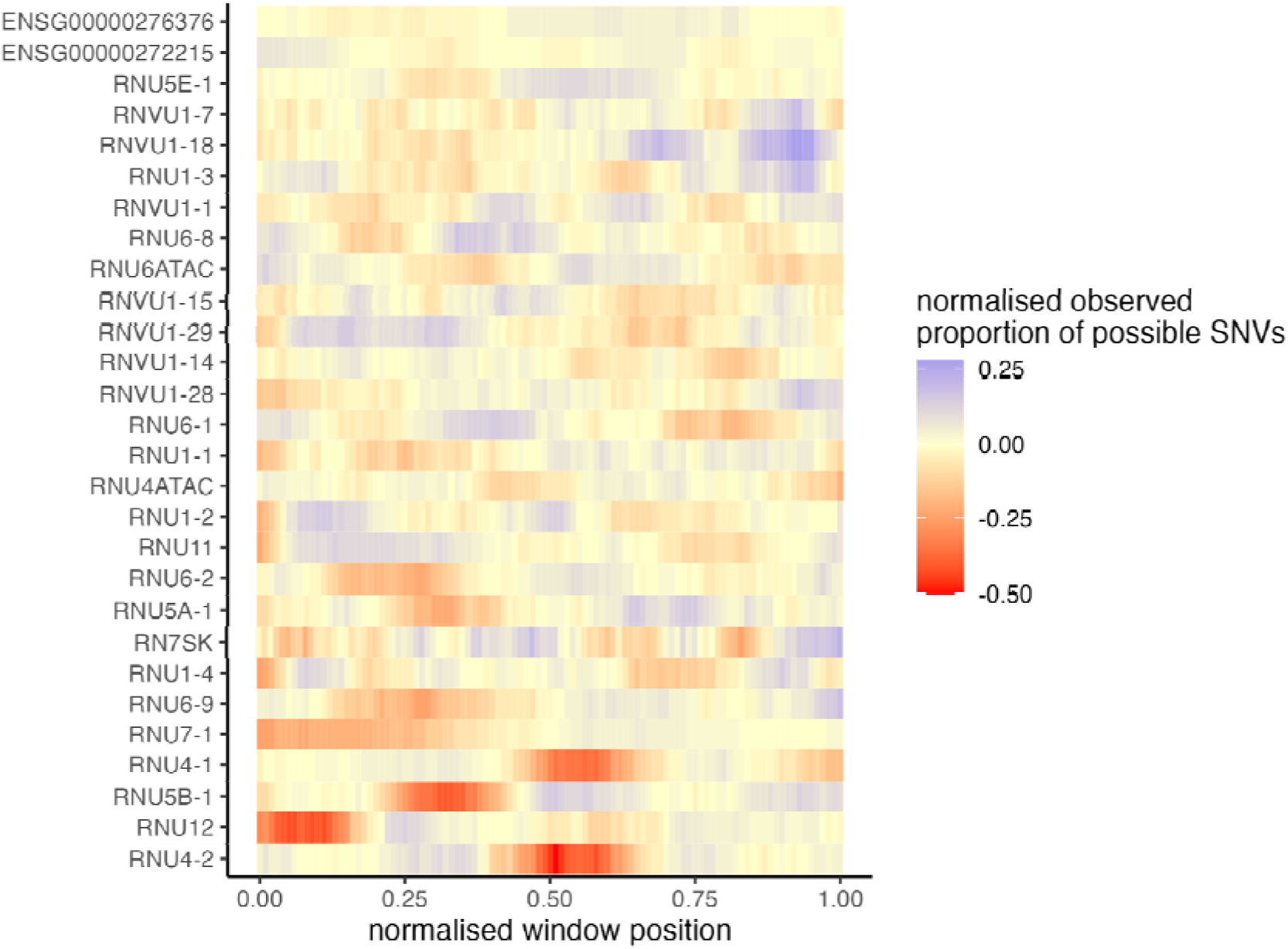
Multiple snRNA genes have regions that are depleted of variation in the population. The proportion of observed SNVs in 490,640 genome sequenced individuals in the UK Biobank, in sliding windows of 18 bp across each snRNA gene, normalised to the median value for each gene.

Finally, we looked for recurrent *de novo* variants in undiagnosed GEL NDD probands that were absent from diagnosed NDD probands, non-NDD probands, and population controls. There are three de novo variants with an allele count ≥3 in the GEL undiagnosed NDD cohort, two in RNU1-2 (chr1:16,895,992:C:T and chr1:16,896,002:A:G), and one in RNVU1-7 (chr1:148,038,767:G:A). However, all three variants are observed at comparable frequencies in non-NDD probands and are also found at relatively high frequencies in population controls (all variants’ AF>0.5% in gnomAD 4.0).

## Discussion

Here, we identified a highly constrained 18 bp region of *RNU4-2* in which variants cause a severe neurodevelopmental phenotype. Variants in this region were identified in 0.68% of individuals with currently undiagnosed NDD in GEL. Assuming a diagnostic rate of 40% upstream of defining our undiagnosed NDD cohort, consistent with recent reports^29^, we estimate that variants in *RNU4-2* could explain 0.41% of all NDD (60/(8841/6*10)). As a comparison, the largest proportion of DD explained by a single gene in a cohort of 31,058 individuals with DD^29^ was 0.47% for *ARID1B*, although we acknowledge that some genes and recognisable syndromes with longstanding associations (e.g., *MECP2*, *SCN1A*, *UBE3A*) will be depleted from this cohort. The proportion of NDD explained by variants in *RNU4-2* would be even higher if restricted to individuals with severe, syndromic NDD. This is consistent with the much lower rate of *RNU4-2* variants in cohorts recruited primarily for autism spectrum disorder (e.g. 3/7,149; 0.042% across SSC^18^, SPARK^37^ and MSSNG^17^).

Our findings underscore the value of large-scale genome sequencing datasets and the importance of considering variants outside of protein-coding regions. This region, despite being within a highly conserved non-coding exon, is not captured by commercially available clinical exome sequencing which primarily captures protein-coding exons^5^. The detailed phenotypic characterisation included here will help prioritise individuals for targeted sequencing of *RNU4-2*.

As *RNU4-2* is a snRNA component of the major spliceosome, we hypothesised that the identified variants would cause a global dysregulation of splicing. However, we did not see this in RNA-seq data derived from blood samples. There are several possible explanations for this result. Firstly, humans have multiple copies of U4-encoding genes, including *RNU4-1* and *RNU4-2*. While *RNU4-2* is the major U4 transcript in the brain, other U4 genes, such as *RNU4-1*, could be expressed at higher levels in blood and play a compensatory role. Future work should look for an effect on splicing in a more relevant cell type or tissue. Secondly, while retention of minor introns is observed in individuals with variants in minor spliceosome components^32^, minor introns represent only a small fraction of all introns across the genome. Large-scale intron retention across major introns would likely be embryonic lethal. The identified variants in *RNU4-2* might therefore have a much more subtle and widespread effect on splicing which is harder to detect. Indeed, variants in U6 snRNA and protein components of the spliceosome situated in the proximity of our *RNU4-2* variants have recently been shown to alter 5’-splice site selection, consistent with this region being involved in subtle regulation of alternative splicing^33,34^.

Finally, given the striking role of *RNU4-2* in NDD we explored whether other snRNA genes could explain undiagnosed cases. We did not find any other snRNAs, or constrained sub-regions of snRNAs, that were enriched for *de novo* variants in NDD cases. We note, however, that these tests have low power given the small size of the genes and regionsmean 139.5 bp and 28.1 bp, respectively). Additionally, we did not explore whether variants in these snRNAs may cause recessive disorders. Variants in the regions we identified should also be investigated in other disease cohorts.

In summary, we identify *RNU4-2* as a novel syndromic NDD gene, explaining ∼0.41% of all individuals with NDD. Including *RNU4-2* in standard clinical workflows will end the diagnostic odyssey for thousands of NDD patients worldwide and pave the way for development of effective treatments for these individuals.

## Methods

### Categorising participants in Genomics England

We defined four groups of individuals in GEL v18. Individuals with NDD (n=13,812) were defined as those with human phenotype ontology (HPO)^35^ and/or International Classification of Diseases 10th Revision (ICD-10) codes^36^ for global developmental delay (HP:0001263, HP:0012736, HP:0011344, HP:0011343, HP:0011342; ICD-10: R62, F80, F81, F82, F83, F88, F89), intellectual disability (HPO: HP:0001249, HP:0002187, HP:0010864, HP:0002342, HP:0001256, HP:0006887, HP:0006889; ICD-10: F70, F71, F72, F73, F78, F79), and/or autism (HPO: HP:0000717, HP:0000729, HP:0000753; ICD-10: F84), or who were recruited to GEL with a normalised specific disease of intellectual disability. NDD individuals were classified as diagnosed (n=3,424) if they were marked as solved or partially solved in the gmc_exit_questionnaire table or had an entry in the submitted_diagnostic_discovery table in GEL Labkey. The remaining 10,388 NDD individuals formed our undiagnosed NDD cohort. Of these, 8,841 are probands. We also identified 21,817 probands without NDD phenotypes (i.e. without the HPO and ICD10 codes detailed above) and 33,122 individuals reported to be unaffected. Our defined cohorts exclude anyone who has subsequently removed consent.

For the majority of our analyses, we used two previously defined datasets within GEL. First, a high-confidence set of *de novo* variants from 13,949 trios^13^. As of 13 March 2024, this dataset includes 12,554 probands with consent: 5,426 probands with undiagnosed NDD, 2,352 with diagnosed NDD, and 4,776 non-NDD probands. *De novo* variants were filtered to those that pass the stringent_filter. Second, an aggregated variant call set (aggV2)^20^ which contains 29,850 probands: 7,519 undiagnosed NDD, 2,903 diagnosed NDD, and 19,428 non-NDD.

### Identifying variants in population datasets

We used data from gnomAD v4.0 (76,215 genome sequenced individuals)^14^, All of Us^15^ (accessed via the publicly available data browser https://databrowser.researchallofus.org/; 245,400 genomes as of 28 March 2023) and the UK Biobank (490,640 genome sequenced individuals)^16^.

### Expanded NDD cohort and clinical data collection

Clinical data were collected from research participants after obtaining written informed consent from the parents or legal guardians, with the study approved by the local regulatory authority. Samples were collected largely through personal communications (NW, AODL, DGM) as variants in this gene have not been prioritised in analysis. On entry into Matchmaker Exchange using the seqr node, one match was made (CD). NW reviewed the National Health Service Genome Medicine Service (NHS GMS; V3) dataset. Samples from NHS GMS were manually checked to remove duplicates with GEL. AODL reviewed the Broad Center for Mendelian Genomics and the GREGoR consortium datasets. DGM contacted additional local collaborators. Clinical data were collected and summarised for features seen across the cohort.

We additionally searched 7,149 trios with autism spectrum disorder and 4,180 sibling control trios from three cohorts: Simons Simplex Collection (SSC; 2,383 cases; 1,938 controls)^18^, SPARK (3,144 cases; 2,190 controls)^37^, and MSSNG (1,622 cases; 52 controls)^17^.

### Generating 1,000 random intergenic sequences

Using the bedtools subtractBed function^38^ we retrieved regions on chromosome 12 that do not overlap with RefSeq transcripts aligned by NCBI. We further removed regions within 10 kbp of an annotated transcript and restricted the remaining regions to those at least 141 bp in length (n=611). We further removed regions overlapping the centromere. We then generated a set of 1,000 random sequences from each intergenic region and then randomly selected 1,000 non-overlapping regions from these.

### Identifying human snRNA genes

We extracted genes with snRNA biotypes from Ensembl genome annotation v111. We filtered out known pseudogenes (i.e. with gene names marked with “P” or identified through manual curation). For each remaining gene, we used BrainVar^28^ RNA-seq expression data to calculate the mean CPM value across the gene. We selected only genes with mean CPM value across all BrainVar samples >5, resulting in a dataset of 28 snRNA genes.

### Assessing variant depletion

Given the high mutability of *RNU4-2* and other snRNA genes, coupled with strong selection pressures on variants, we did not think that conventional mutational models would be well calibrated to assess variant depletion. Instead, we devised a sliding window-based strategy to identify regions within snRNA genes that are relatively depleted of SNVs. We split genes into 18 bp sliding windows (chosen as it is the size of the region defined in *RNU4-2*) and tallied the number of SNVs observed in UK Biobank 500k genome sequencing data within that window, divided by the total number of possible SNVs (i.e. 18x3). The proportion of possible SNVs observed in each window was normalised to the median across all sliding windows in that gene (i.e. the per-gene median proportion observed was subtracted from each value). Depleted regions were defined as those spanning windows with a deviation from the per-gene median of at least 20%, i.e. normalised observed proportion of possible SNVs < −0.2. The same calculation was performed on 1,000 randomly selected 141 bp intergenic regions on chr12 (see above). A one-way approximative (Monte Carlo) Fisher-Pitman test was conducted to show the median observed proportion of possible SNVs was significantly higher for *RNU4-1* and *RNU4-2* compared to the distribution in the 1,000 random regions.

### RNA-sequencing of individuals with RNU4-2 variants

Blood was collected from a subset of 100,000 Genomes Project probands in PaxGene tubes to preserve RNA at the time of recruitment. RNA was extracted, depleted of globin and ribosomal RNAs, and subjected to sequencing by Illumina using 100 bp paired-end reads, with a mean of 102M mapped reads per individual. Alignment was performed using Illumina’s DRAGEN pipeline. FRASER2^26^ and OUTRIDER^25^ were used to detect abnormal splicing events and expression differences with samples run in batches of 500, both run via the DROP pipeline^39^.

### Analysing RNU4-2 and RNU4-1 expression

We used the BrainVar^28^ dataset to assess patterns of whole-gene expression of *RNU4-2* and *RNU4-1* in the human cortex across prenatal and postnatal development. This dataset includes bulk-tissue RNA-seq data from 176 de-identified postmortem samples of the dorsolateral prefrontal cortex (DLPFC, n=167 older than 10 post-conception weeks) or frontal cerebral wall (n=9 younger than 10 post-conception weeks), ranging from 6 post-conception weeks to 20 years of age. The 100 bp paired-read RNA-seq data from BrainVar were aligned to the GRCh38.p12 human genome using STAR aligner^40^, and gene-level read counts for GENCODE v31 human gene definitions were calculated with DEXSeq^41^ and normalised to counts per million (CPM)^42^.

### Prenatal prefrontal cortex ATAC-seq data

Methods of generating ATAC-seq have been described previously^43^, which is the source of the data shown here. Briefly, fresh prenatal (18 and 19 gestational weeks) brain samples were dissected within 2 hours of elective termination to extract the entire telencephalic wall, from the ventricular zone to the meninges. Intact nuclei were isolated by manually douncing the tissue on ice using a loose pestle douncer then lysed on ice for 10 minutes by adding a solution with 0.1% NP-40. Nuclei were spun down by centrifugation then resuspended and exposed to Tagmentation Enzyme for 30 minutes at 37C. The ATAC-seq library was generated using Illumina barcode oligos, amplified by high-fidelity PCR, and sequenced on the Illumina HiSeq 2500 using paired-end sequencing. Reads were aligned to GRCh38 using the ENCODE ATAC-seq pipeline with default parameters^44^. A UCSC Browser track of per nucleotide ATAC-seq counts was used to assess the region around *RNU4-2* and *RNU4-1*.

### Burden testing and statistical analysis

The enrichment of *de novo* variants across each of 28 snRNA genes and 14 constrained sub-regions was assessed in undiagnosed NDD probands compared to non-NDD probands using the high-confidence *de novo* callset. Odds ratios and associated *P*-values were calculated using Fisher’s exact test in R. A *P*-value threshold of 0.0031 was used to assess statistical significance as a Bonferroni correction accounting for 16 tests.

## Data Availability

All data produced in the present work are contained in the manuscript.

## Acknowledgements

We would like to thank Peter O’Donovan, Mitra Sato, Mimoza Hoti and Joanne Yang from Genomics England for their help with clinician collaboration and Airlock requests.

YC is supported by a studentship from Novo Nordisk. NW is supported by a Sir Henry Dale Fellowship jointly funded by the Wellcome Trust and the Royal Society (220134/Z/20/Z). GL was supported by the Fonds de recherche en santé du Québec (FRQS), VSG by NIAMS K23AR083505, and SLS by a fellowship from Manton Center for Orphan Disease Research at Boston Children’s Hospital. The research was supported by grant funding from Novo Nordisk and the Rosetrees Trust (PGL19-2/10025 to NW), the Simons Foundation Autism Research Initiative (SFARI #736613, SJS), the NIMH (R01 MH129751 to SJS), the HDR-UK Molecules to Health Records Driver Programme (SJS), the Australian National Health and Medical Research Council (1164479, 1155244, GNT2001513), the Australian NHMRC Centre for Research Excellence in Neurocognitive Disorders (NHMRC-RG172296), the Australian Medical Research Future Fund (MRF2007677, GHFM76747), NHGRI (U01HG011762, U01HG011745, U24HG011746, UM1HG008900, U01HG011755, R21HG012397, and R01HG009141), NINDS (U01NS134358, U01NS106845, U54NS115052, 1U24NS131172), the Chan Zuckerberg Initiative Donor-Advised Fund at the Silicon Valley Community Foundation (funder DOI 10.13039/100014989) grants 2019-199278, 2020-224274, (https://doi.org/10.37921/236582yuakxy), the US Department of Defense Congressionally Directed Medical Research Programs (PR170396), the National Institute of Neurological Disorders and Stroke of the National Institutes of Health (U01HG007709, U01HG007942, and U01HG010217), and the Clinical Translational Core of the Baylor College of Medicine IDDRC (P50HD103555) from the Eunice Kennedy Shriver National Institute of Child Health and Human Development. The content is solely the responsibility of the authors and does not necessarily represent the official views of the Eunice Kennedy Shriver National Institute of Child Health and Human Development or the National Institutes of Health. The Rare Disease Flagship acknowledges financial support from the Royal Children’s Hospital Foundation, the Murdoch Children’s Research Institute and the Harbig Foundation. Massimo’s Mission acknowledges funding support from the Australian Government Department of Health and Aged Care (EPCD000034). Sequencing and analysis were supported by the Deutsche Forschungsgemeinschaft (DFG) Research Infrastructure West German Genome Center (project 407493903) as part of the Next Generation Sequencing Competence Network (project 423957469). Short-read genome sequencing was carried out at the production site Cologne (Cologne Center for Genomics; CCG). CD received the DFG 458099954 as part of the DFG Sequencing call #3. We also thank Sabine Kaya for technical assistance, Christopher Schröder for bioinformatic analysis. SMS is supported by the Epilepsy Society.

This research was made possible through access to data in the National Genomic Research Library, which is managed by Genomics England Limited (a wholly owned company of the Department of Health and Social Care). The National Genomic Research Library holds data provided by patients and collected by the NHS as part of their care and data collected as part of their participation in research. The National Genomic Research Library is funded by the National Institute for Health Research and NHS England. The Wellcome Trust, Cancer Research UK and the Medical Research Council have also funded research infrastructure.

## Supplementary Figures

**Supplementary Figure 1:**
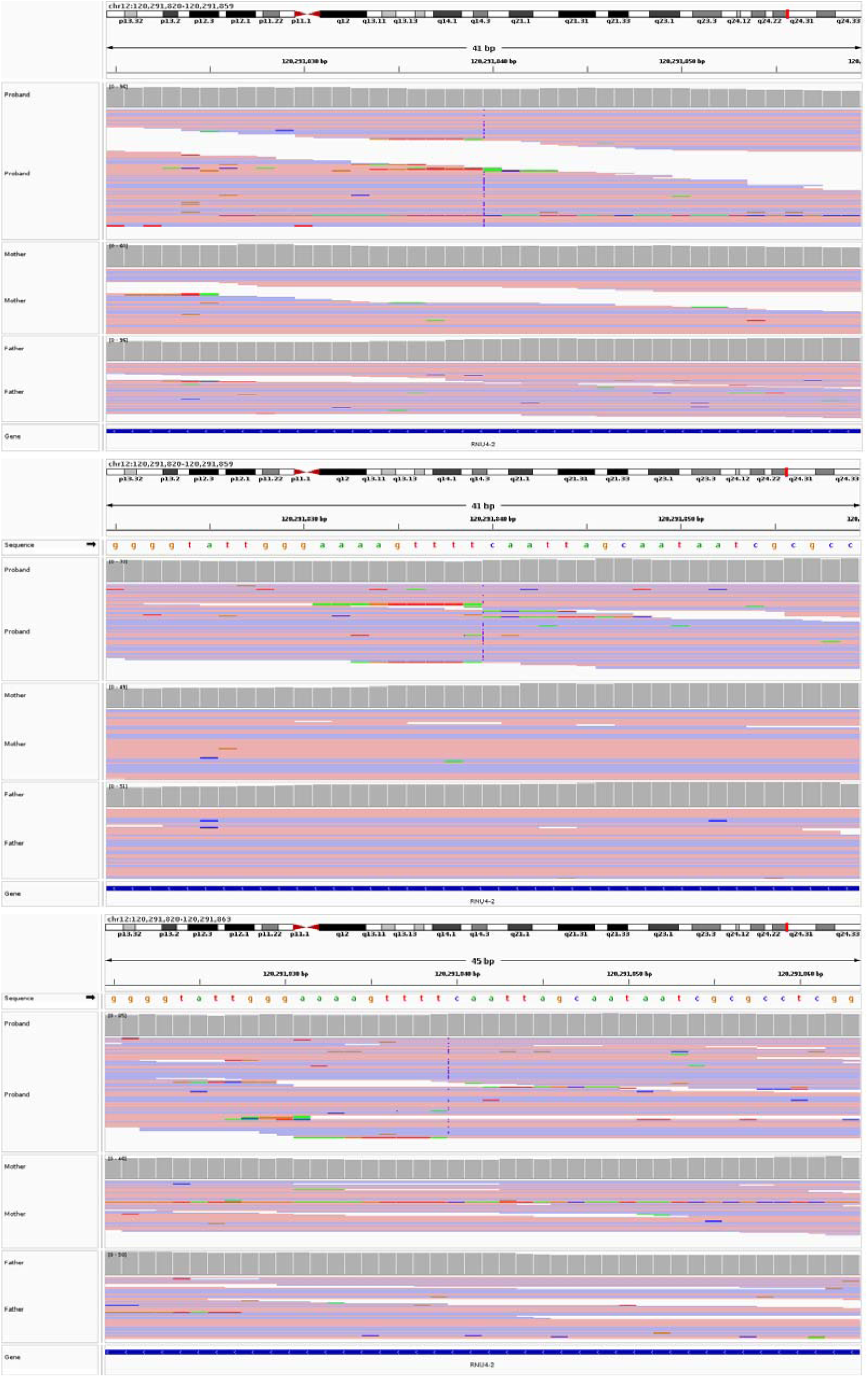
Example IGV plots of the region surrounding the n.64_65insT variant in three trios.

**Supplementary Figure 2:**
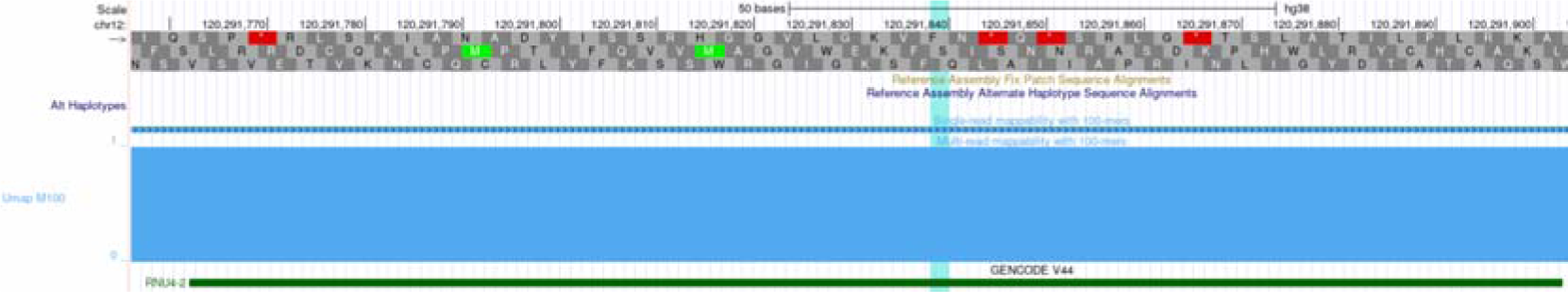
Screenshot from the UCSC Genome Browser showing high mappability for 100-mers across the *RNU4-2* gene.

**Supplementary Figure 3:**
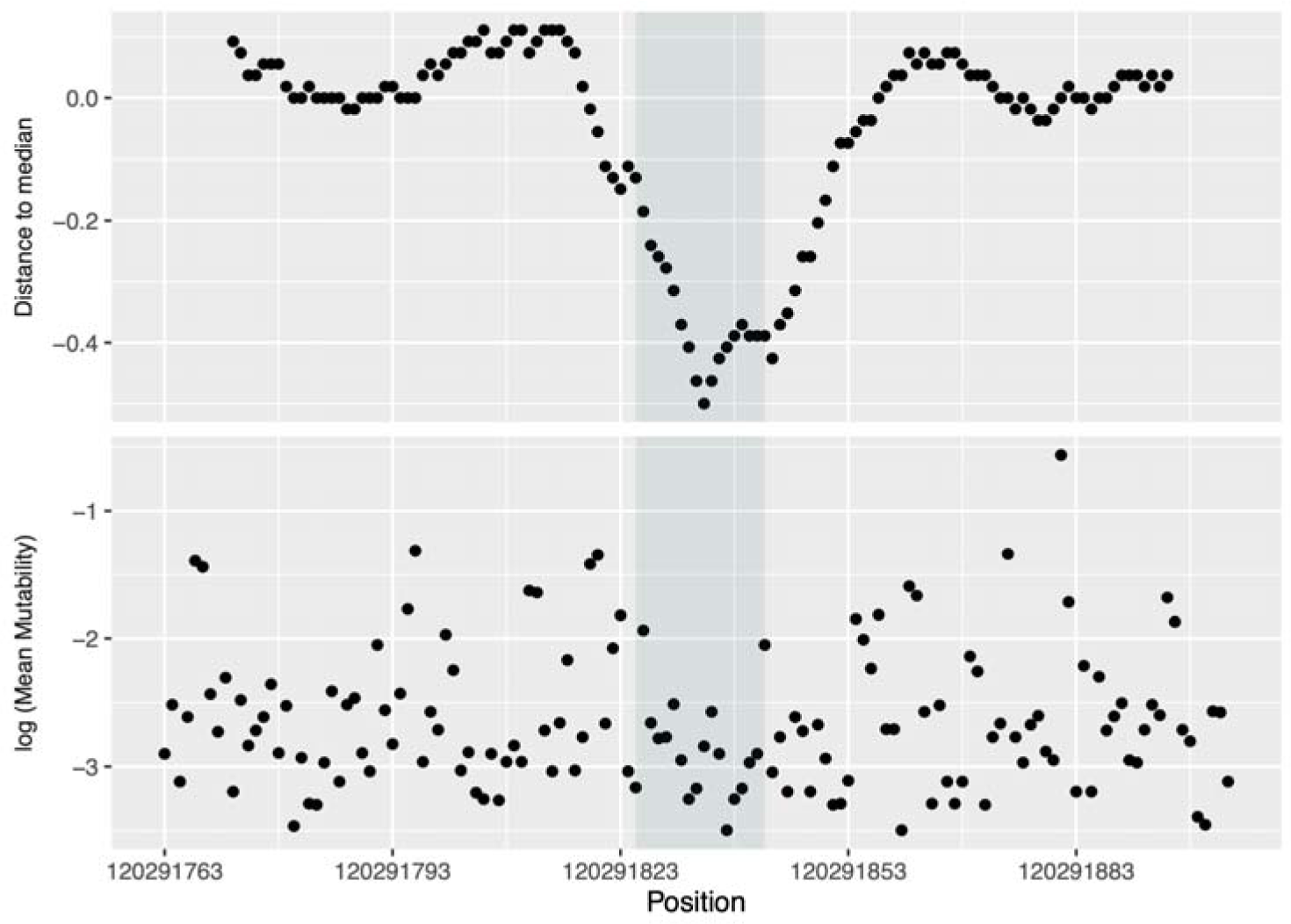
(top) Distance to the median proportion of all possible SNVs that are observed in the UK Biobank in 18 bp sliding windows across the length of RNU4-2. A clear region of depletion compared to the rest of the gene is observed in the centre. (bottom) Log transformation of the mean Roulette^45^ mutability across the 3 possible SNVs within a site.

**Supplementary Figure 4:**
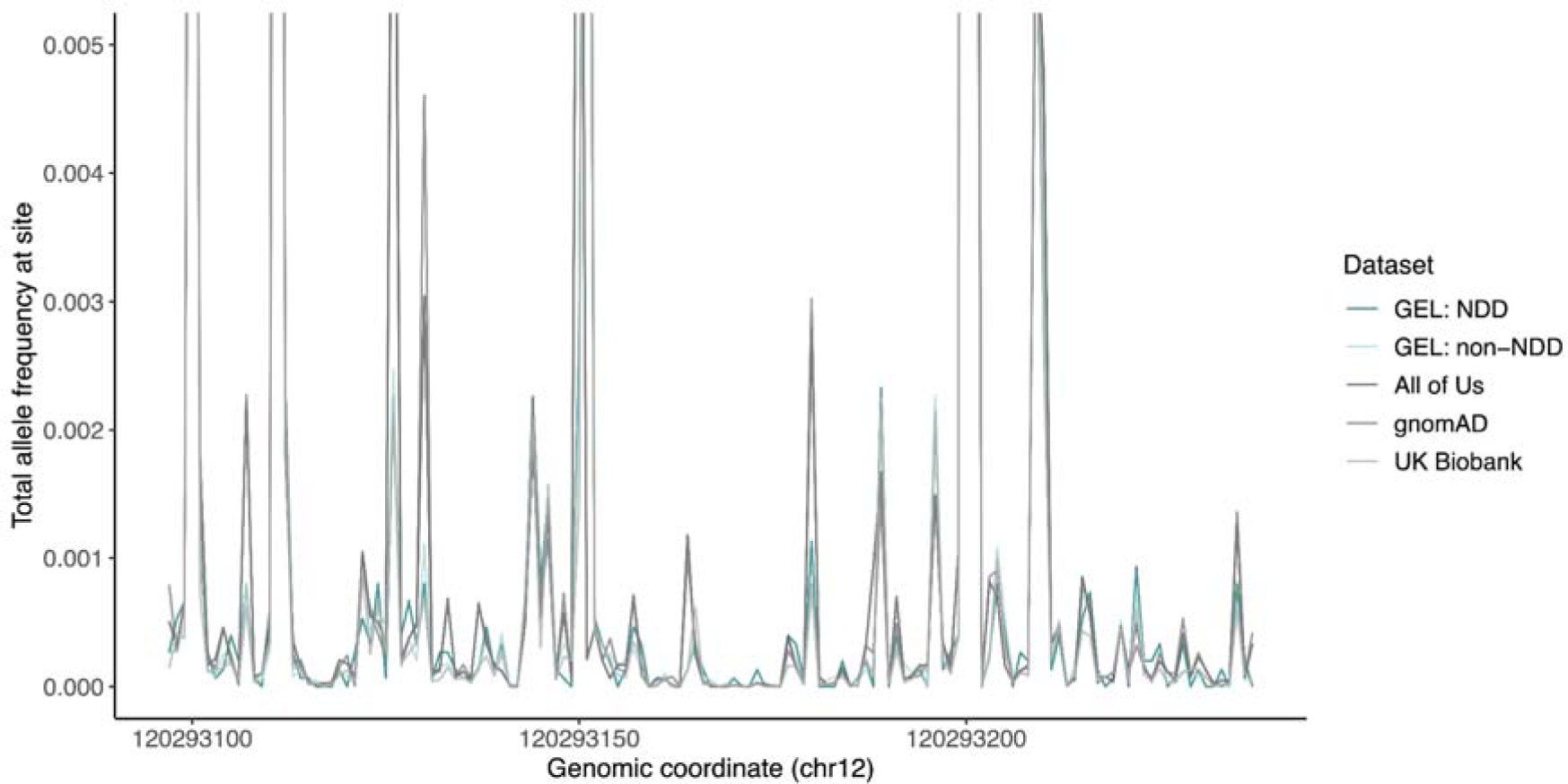
Total allele frequency at each site of *RNU4-1* in five datasets. In contrast to *RNU4-2* (Figure 2a), variants in *RNU4-1* have higher allele frequencies. A similar region of depletion is seen in the centre of *RNU4-1* (quantified in Figure 4), but this is not enriched for variants in GEL NDD or non-NDD individuals.

**Supplementary Figure 5:**
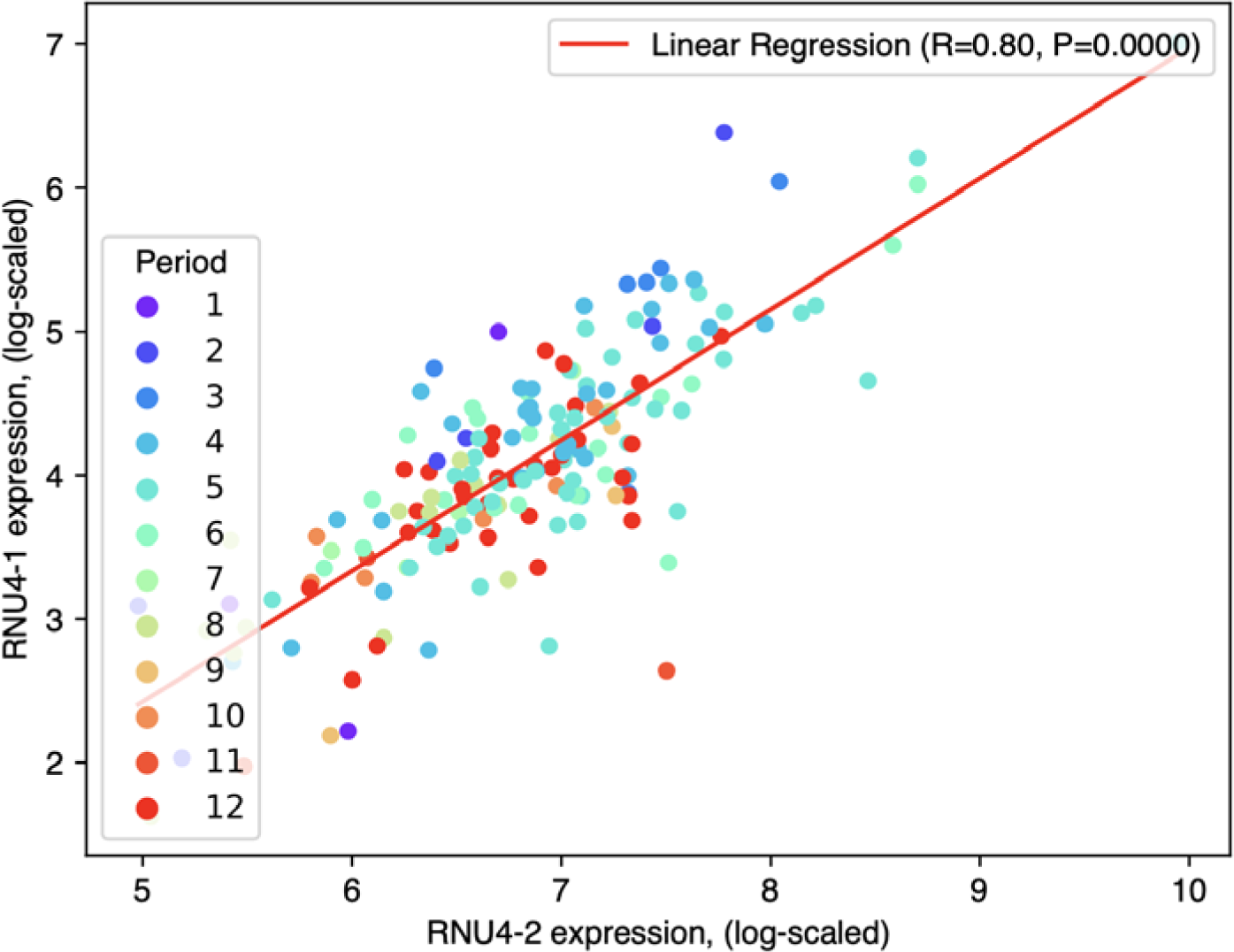
Correlation between *RNU4-1* and *RNU4-2* expression in RNA-seq data from human cortex across prenatal and postnatal development from BrainVar^28^.

**Supplementary Figure 6:**
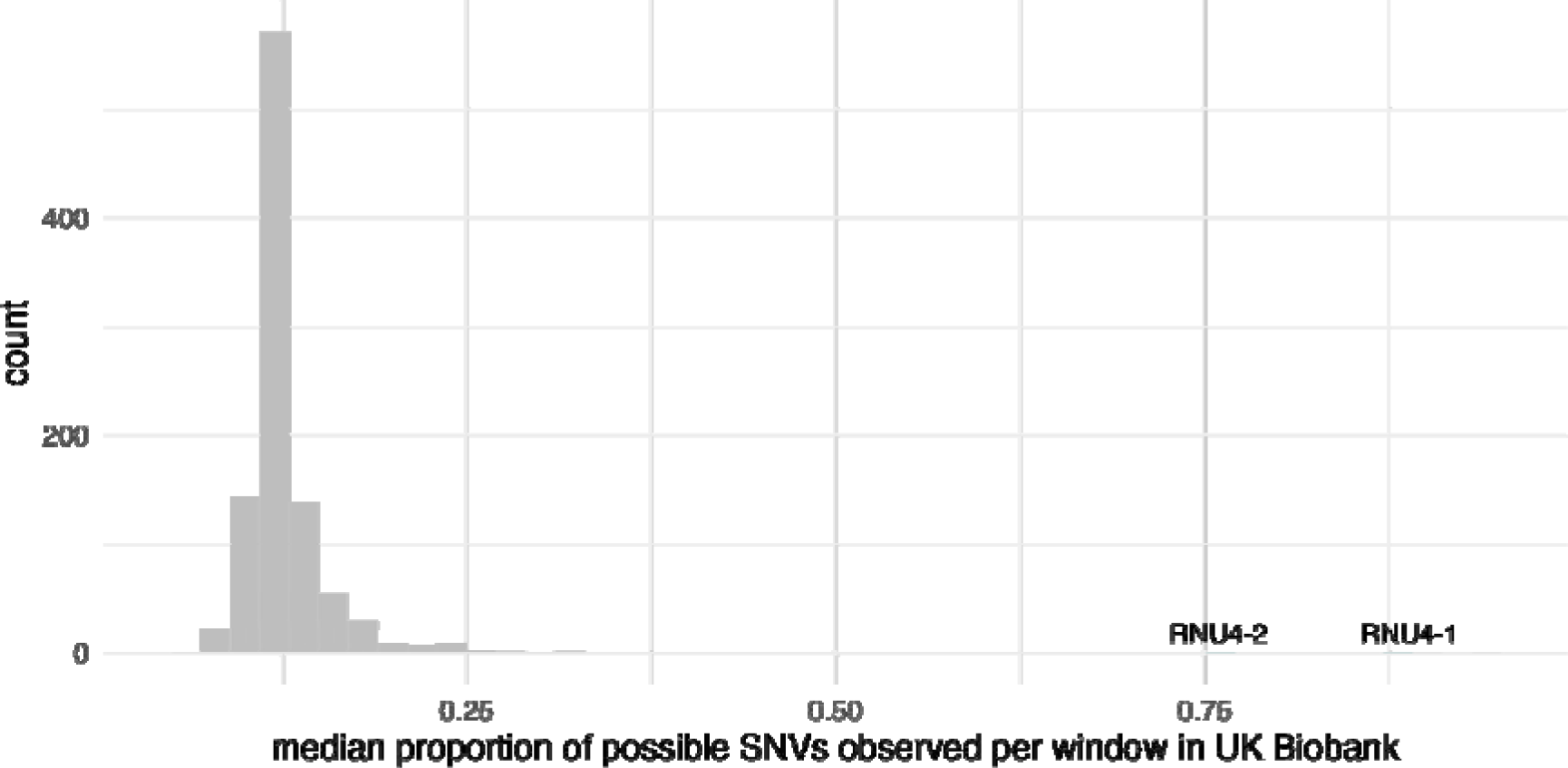
Median proportion of possible SNVs observed in UK Biobank per 18 bp window across 1,000 intergenic regions on chromosome 12 (grey) and *RNU4-1*, *RNU4-2* (teal).

**Supplementary Figure 7:**
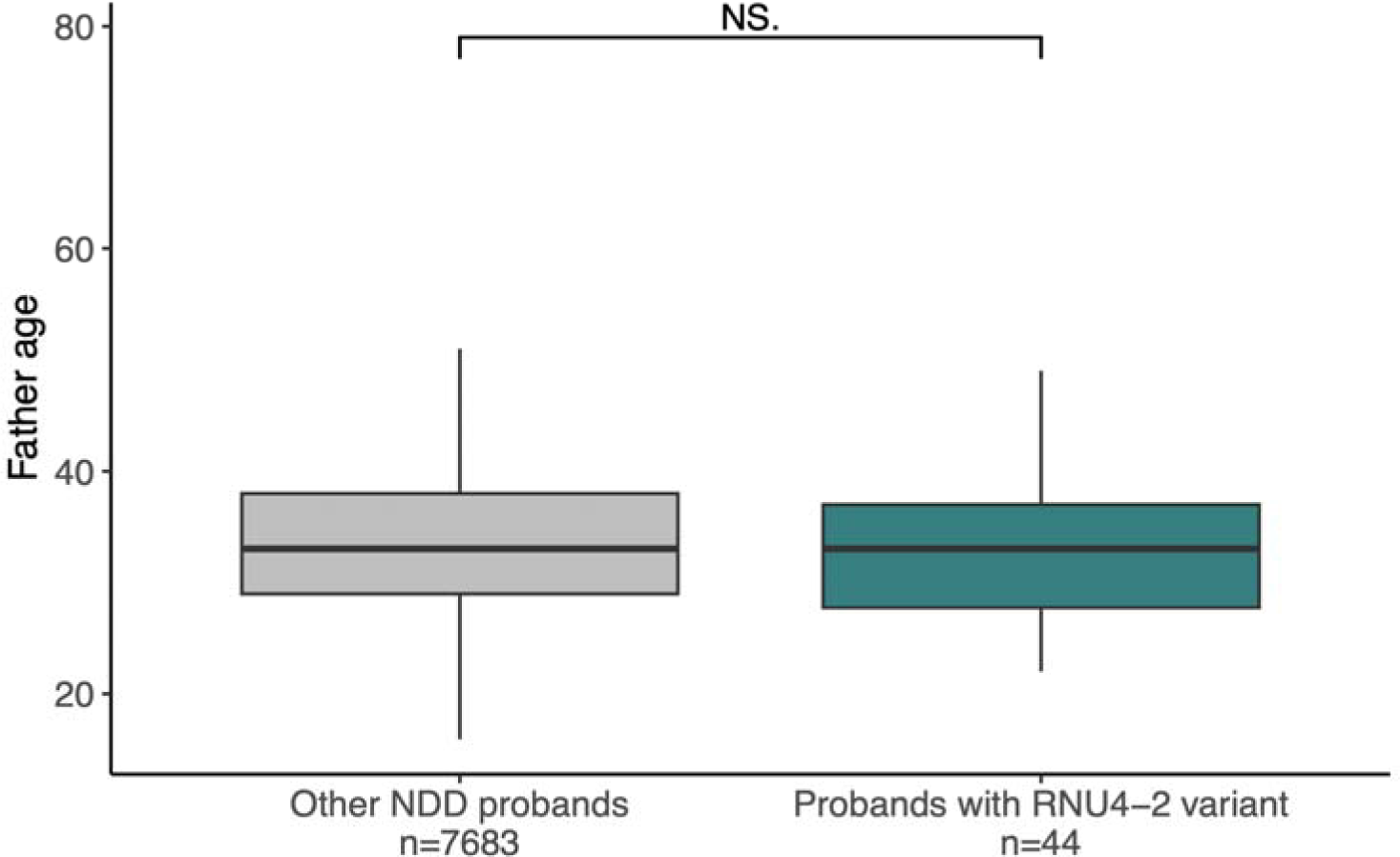
Comparison of paternal age for probands with fathers recruited into GEL.

## Supplementary Tables

**Supplementary Table 1**: The number of probands with the n.64_65insT variant and all other individuals with NDD with HPO terms corresponding to phenotypes observed in ≥ 5 individuals compared to all other NDD probands. These data are plotted in Figure 1a. A P-value threshold of 2.94×10^-3^ was used to assess statistical significance (Bonferroni adjusted for 17 tests).

**Supplementary Table 2**: ICD10 and ICD9 codes for individuals with single base pair insertions between codons 64 and 65 of *RNU4-2* and *RNU4-1* in the UK Biobank.

**Supplementary Table 3**: Outliers predicted by OUTRIDER and FRASER2 in RNA-seq data for five individuals with *RNU4-2* variants compared to 5,409 controls. A P-value threshold of 0.017 was used to assess statistical significance (Bonferroni adjusted for 3 tests).

**Supplementary Table 4**: Detailed clinical information for 25 individuals with RNU4-2 variants. SNVs are highlighted in pink, and the individual with an alternate indel in blue. Blank spaces indicate that data were not provided.

**Supplementary Table 5**: Detailed phenotypic information for individuals with the n.64_65insT variant across cohorts.

**Supplementary Table 6**: Mean expression of U4 genes in prefrontal cortex across all samples in BrainVar.

**Supplementary Table 7**: Genomic coordinates of, and burden testing results for snRNA genes.

**Supplementary Table 8**: Sub-regions of snRNA genes identified as depleted of variation and burden testing results in these regions.

